# Clinical mechanism of ribavirin action in Hepatitis C treatment: insights from the STOPHCV-1 randomised trial

**DOI:** 10.64898/2026.04.14.26350846

**Authors:** Mahdi Moradi Marjaneh, Anjna Badhan, Haiting Chai, Olivia Hadfield, Yixuan Chen, Ziping Wang, Emma C Thomson, Graham P Taylor, A Sarah Walker, M Azim Ansari, Eleanor Barnes, Graham S Cooke

**Author notes:** Correspondence to: Prof Graham S Cooke, Imperial College London, South Kensington Campus, London SW7 2AZ, UK. These authors contributed equally to this work (Mahdi Moradi Marjaneh, Anjna Badhan).

## Abstract

**Background:** Ribavirin is a guanosine analogue with clinical antiviral activity against a range of RNA viruses including hepatitis C virus (HCV), respiratory syncytial virus and Lassa virus. Several potential mechanisms of action have been proposed, but there is limited data supporting them clinically.

**Methods:** We studied 196 HCV-infected participants from a trial of short-course directly antiviral therapy (STOPHCV-1) which included a factorial randomisation to ribavirin versus no ribavirin. Deep sequencing of the HCV genome was performed on samples with detectable viremia from three time-points: baseline (n = 191), day 3 of treatment (n = 25) and post-treatment failure (n = 47).

**Results:** Ribavirin exposure significantly increased total mutational load at treatment failure (*P* = 0.0065) and enriched classical ribavirin-associated transitions, including G→A (*P* = 0.026) and C→U (*P* = 0.004), along with other key changes including A→G (*P* = 0.005), U→C (*P =* 0.023), C→G (*P =* 0.010), and U→A (*P =* 0.026). Ribavirin’s mutational signature was broad, not dominated by G-related changes. Region-specific analyses demonstrated this increase was broadly distributed across the viral genome, without strong evidence for protection of specific regions. Non-synonymous to synonymous mutation ratios (dN/dS) rose at day 3 (*P* = 5.5e-5) before declining at failure (*P* = 8.5e-7), with trends toward higher dN/dS in the ribavirin group at day 3 (*P* = 0.06).

**Conclusions:** Ribavirin acts as a potent *in vivo* mutagen, driving viral populations toward genome-wide diversity rather than selecting a few highly fit drug-resistant clones. These findings support an error-catastrophe model.

## Introduction

Ribavirin, a synthetic guanosine nucleoside analogue, has been used for over four decades as a broad-spectrum antiviral. It has clinical activity against a number of RNA viruses, most notably hepatitis C virus (HCV). Advances in directly acting antiviral (DAA) therapy for HCV mean ribavirin is no longer routinely recommended for treatment but it remains a treatment option for respiratory syncytial virus (RSV), influenza and some viral haemorrhagic fever viruses (e.g. Lassa) (Graci & Cameron, 2006; McClung et al., 1983; Thomas et al., 2012).

Despite its longevity in clinical use and importance in historical HCV therapy, the molecular mechanisms underpinning its antiviral activity remain incompletely understood and, in some cases, debated.

Several, often overlapping, mechanisms have been proposed to explain ribavirin’s antiviral effects, including inhibition of viral polymerase activity, depletion of intracellular guanosine triphosphate through inosine monophosphate dehydrogenase (IMPDH) inhibition, lethal mutagenesis, and modulation of host immune responses (Beaucourt & Vignuzzi, 2014; Crotty et al., 2000; Graci & Cameron, 2006). Despite these proposed roles, no single pathway fully accounts for its broad antiviral efficacy across diverse RNA viruses.

Among the proposed mechanisms, lethal mutagenesis (error catastrophe) has emerged as one of the most compelling hypotheses (Graci & Cameron, 2006; Thomas et al., 2012). This model suggests that ribavirin elevates the viral mutation rate by ambiguous base-pairing during RNA synthesis, producing characteristic transition mutations such as G→A and C→U. Once mutation levels surpass the virus’s natural error threshold, ongoing replication becomes unstable, leading to reduced fitness and eventual viral collapse. Experimental studies *in vitro* have demonstrated this mutagenic effect in several RNA viruses, including poliovirus, influenza, SARS-CoV-2, and HCV (Crotty et al., 2000; Martinez-Gonzalez et al., 2024; Somovilla et al., 2023). Yet, direct clinical evidence supporting this mechanism *in vivo* has been limited.

HCV provides a valuable model for investigating ribavirin’s mutagenic potential due to its high replication rate, error-prone RNA-dependent RNA polymerase (RdRp), and diverse quasispecies structure. Prior *in vitro* work has shown that ribavirin exposure increases the frequency of transition mutations in the NS3 and NS5B genes and may alter the quasispecies distribution of viral populations (Mejer et al., 2020; Nakamura et al., 2022; Saito et al., 2020). However, most clinical studies have lacked either randomisation, longitudinal sampling, or deep sequencing capable of capturing low-frequency variants. As a result, the true extent of ribavirin-induced mutagenesis and its influence on viral evolution in patients have remained unclear.

The STOPHCV-1 clinical trial provides a unique opportunity to address these gaps (Cooke et al., 2021). This large, randomised, multicentre study of short-course DAA therapy included a factorial randomisation to ribavirin versus no ribavirin, generating the largest clinical dataset available for analysing ribavirin’s genomic effects. By combining deep sequencing of baseline, on-treatment (Day 3), and post-treatment failure samples, we aimed to characterise ribavirin’s impact on mutation rate, substitution spectrum, and selective pressure over the course of therapy.

## Methods

### Study design and participants

STOPHCV-1 was a multicentre, randomised, open-label trial conducted across 14 UK NHS Hospital Trusts between March 2016 and August 2018. The trial enrolled 202 adults aged ≥18 years with chronic HCV genotypes 1a, 1b, or 3 infection of at least 6 months’ duration. Participants were randomly assigned to two short-course treatment approaches, a fixed 8 weeks duration vs 4-7 weeks duration depending on baseline HCV viral load (Cooke et al., 2021). DAAs were predominantly ombitasvir, paritaprevir, ritonavir with or without dasabuvir depending on genotype. A factorial randomisation allocated participants to receive or not receive ribavirin with their first-line DAA therapy. Ribavirin dosing was weight based twice daily (<75kg 1000mg/day, ≥75kg 1200mg/day). Treatment success was assessed by sustained virological response at 12 weeks post-therapy (SVR12).

### Baseline and on-treatment comparisons

Baseline demographic, clinical, and laboratory characteristics were compared between the ribavirin and no-ribavirin groups using standard statistical tests. Continuous variables were summarised as means with standard deviations and compared using two-sample t-tests or Wilcoxon rank-sum tests when normality assumptions were not met. Categorical variables were compared using χ^2^ tests or Fisher’s exact tests when expected cell counts were <5. All tests were two-sided, and *p* < 0.05 was considered statistically significant.

On-treatment virological measures (HCV viral load at days 3, 7, and 14, and absolute and relative viral load declines from day 0-3) were derived from trial viral load measurements performed locally at participating NHS Trusts. A variety of assays were used, including Cobas Amplicor v2, Abbott RealTime, Aptima QuantDX, and Roche CAP-CTM, each with assay-specific lower limits of detection (LOD). Values reported as below the assay detection threshold were identified using the accompanying result type variable and treated as left-censored observations, with the lower bound set to zero and the upper bound set to the assay-specific LOD. To account for interval censoring and heterogeneity in assay sensitivity, viral load measurements were analysed on the log_10_ scale using interval regression models (Gaussian distribution), implemented in R using the *survival* package (v3.8.3). Early viral decline (day 0-3) was analysed similarly using interval-censored differences between baseline and day 3 measurements. Relative declines were summarised using midpoint estimates.

### Sequencing

Baseline and post-treatment failure samples were assayed as previously described (Lin et al., 2021). Day 3 plasma samples were retrieved from the HCV Research UK Biobank (http://www.hcvresearchuk.org/) and assayed for this specific study. HCV viral loads were quantified using the COBAS TaqMan PCR assay (Roche) according to the manufacturer’s instructions, and samples with viral loads >500 IU/mL were included. Viral RNA was extracted from 1 mL of plasma using the QIAamp® Viral RNA Mini Kit (Qiagen), as described previously (Thomson et al., 2016). Samples were sequenced using two complementary approaches.

#### 1. Target enrichment whole-genome sequencing

Whole-genome sequencing was performed using the SureSelect XT HS2 RNA System on the Magnis NGS Prep platform (Agilent), following the manufacturer’s protocol. Briefly, 10 ng of RNA input was used with 14 pre-capture and 22 post-capture PCR cycles. HCV-specific probes were used for hybridization and capture (SureSelect XT HS2 RNA and SureSelect XT Community Design Pan-HCV; Agilent, Cat#5191-6709). Library size and quality were assessed using the 4200 TapeStation (D1000 kit; Agilent), and libraries were quantified using Qubit dsDNA Broad Range and High Sensitivity Assay kits (Thermo Fisher Scientific). Semi-quantitative assessment of HCV content was performed by qPCR using the KAPA SYBR Fast kit (KAPA Biosystems) with HCV-specific primers (Davalieva et al., 2014). Libraries were pooled in equimolar proportions, normalized, and sequenced on the Illumina iSeq 100 platform using 2×100 bp paired-end reads.

#### 2. Pre-amplification PCR sequencing

Targeted sequencing of HCV regions (E1, E2, NS3, NS4B, and NS5B) was performed using genotype-specific primers in five or six overlapping amplicons per genotype/subtype, as previously described (Thomson et al., 2016). Viral RNA was amplified via single-step reverse transcription PCR (SuperScript III reverse transcriptase; Invitrogen), followed by nested PCR. PCR products were purified using the QIAquick kit (Qiagen), pooled to include alternate amplicons, and quantified using Qubit assays prior to library preparation with the Nextera XT DNA Sample Preparation Kit (Illumina). Indexed libraries were sequenced on the Illumina MiSeq platform using the v2 deep sequencing reagent kit.

### Sequence processing and variant calling

#### Read processing and quality control

Raw paired-end FASTQ files underwent initial quality control using FastQC (v0.11.7) (Andrews, 2010). Read preprocessing was performed using a custom wrapper built around the preprocessReads function from QuasR (v1.36.0), which trimmed low-quality bases and removed reads shorter than 14 nucleotides. Each sample was additionally processed with Trim Galore (v0.6.7) for adapter trimming (Krueger, 2015).

#### Human read removal

Pre-processed paired-end reads were aligned against the human reference genome (GRCh38) using Bowtie2 (v2.2.9) (Langmead & Salzberg, 2012), and all mapped reads were discarded. Only read pairs for which both mates remained unmapped were retained for downstream analyses.

#### Genotyping and reference assignment

To assign each sample to its closest HCV reference genome, host-filtered FASTQ files were converted to FASTA format and queried against the full HCV reference genome database (https://hcv.lanl.gov/) using blastn from blast+ (v2.11.1) (Camacho et al., 2009). BLAST hits were sorted by read and alignment score, and the top five hits per read were retained. Read-level alignments were aggregated to quantify how many reads supported each candidate reference genome, and the genome with the highest support was selected as the best-matching reference for that sample.

#### Read alignment

Host-filtered reads were then aligned to the selected HCV reference genome using Bowtie2 (v2.2.9) (Langmead & Salzberg, 2012). Concordantly aligned read pairs were extracted from each mapped BAM file using Samtools (v1.16) (Li et al., 2009), generating cleaned BAM files containing only properly paired viral reads.

#### Variant calling

Variant identification was performed using quasitools (v0.7.0) (Marinier et al., 2019). ntvar was used to detect nucleotide-level variants, codonvar to call codon-level variants, and aavar to identify amino-acid variants, applying a minimum amino-acid variant frequency threshold of 5%. Amino-acid coverage across the HCV genome was computed using aacoverage.

#### Diversity analysis (sample-level)

Intra-host viral diversity was quantified using the complexity module from quasitools (v0.7.0), which computes per-position measures of haplotype counts, Shannon entropy, and the Gini-Simpson index across the viral genome. Shannon entropy and the Gini-Simpson index quantify viral population diversity based on the distribution of variant or haplotype frequencies. Shannon entropy measures the uncertainty in the system, calculated as −∑p_i ln(p_i), while the Gini-Simpson index reflects the probability that two randomly selected viral genomes differ, calculated as 1 − ∑p_i^2^, where p_i represents the frequency of each variant or haplotype. Sample-level diversity metrics were obtained by averaging these per-position values across all genomic sites (excluding positions with missing values). Statistical analyses were adjusted for sequencing depth to account for potential coverage-related biases.

### Cohort-level and statistical analyses

All downstream analyses were conducted in R using custom scripts. Data from all samples were merged into unified analytical datasets and linked with sample- and participant-level metadata.

#### Mutation frequency analyses

Quasitools outputs were processed using dplyr (v1.1.4), tidyr (v1.3.1), readr (v2.1.5), and stringr (v1.5.1) to derive per-sample mutation profiles (overall and by substitution type). Mutations were filtered for ≥100× coverage. Covered viral genome length was defined as the number of genomic positions with sequencing depth ≥100× for each sample. Mutation counts were normalised by the number of covered positions to obtain persite mutation frequencies, thereby accounting for differences in genome coverage between samples. Group comparisons between ribavirin and non-ribavirin groups were performed using Wilcoxon rank-sum tests (rstatix v0.7.2) and visualised with violin plots and boxplots (ggplot2 v3.5.2).

#### Depth-adjusted modelling

To assess whether sequencing depth influenced mutation-frequency estimates or ribavirin comparisons, beta-binomial regression models were fitted using glmmTMB (v1.1.13). Alternate and reference counts from quasitools ntvar VCFs were merged with metadata. Two models were fitted per time point: (i) a base model including ribavirin status, and (ii) a depth-adjusted model additionally including site-specific coverage as a fixed-effect covariate. Models were compared using likelihood-ratio tests, and changes in the ribavirin coefficient (Δ-estimate = β_with-depth - β_no-depth) quantified depth effects.

#### Diversity analysis (cohort-level)

Quasitools complexity metrics were aggregated using dplyr (v1.1.4) and compared across ribavirin groups and time points using Wilcoxon rank-sum tests and, when relevant, linear models adjusting for the continuous gap covariate. Diversity trajectories were visualised using line, violin, and boxplots (ggplot2 v3.5.2).

#### Evolutionary analysis

Gene-wise dN/dS ratios were computed within each sample using quasitools by aggregating nonsynonymous and synonymous variants across coding regions. These gene-level estimates of within-host selective pressure were compared between treatment groups (Ribavirin vs Non-Ribavirin) using Wilcoxon rank-sum tests implemented in rstatix (v0.7.2), and visualised using ggplot2 (v3.5.2).

## Code availability

All scripts used for the analyses in this study are available at:

https://github.com/MahdiMoradiMarjaneh/Ribavirin_STOPHCV1

### Results

### Participant characteristics by ribavirin use

In total, 202 participants were randomised; however, four were excluded due to the absence of stored baseline samples, and two were excluded due to HCV genotype 3 infection to avoid bias arising from the small subgroup size. Consequently, 196 participants were included in this analysis, including 98 ribavirin recipients.

Baseline characteristics were generally well balanced between ribavirin and no-ribavirin groups, as expected by the randomisation (**Table 1**). The two groups were comparable in age (45.1 vs 46.3 years, *P* = 0.41), sex (34.7% vs 27.6% female, *P* = 0.28), baseline HCV viral load (5.61 vs 5.83 log_10_ IU/mL, *P* = 0.09), and HCV subtype distribution (84.7% vs 81.6% genotype 1a, *P* = 0.57). The prevalence of the IL28B CC genotype was similar between groups (29.8% vs 33.0%, *P* = 0.64), as was the proportion with CT or TT genotypes (70.2% vs 67.0%). Liver stiffness measurement, liver biochemistry, renal function, and the prevalence of HIV co-infection, depression, or illicit substance use also did not differ significantly. The only significant baseline difference was in BMI, which was lower in the ribavirin group (24.3 vs 26.5 kg/m^2^, *P* < 0.001). Of note, 18 factors were tested for this analysis, meaning one would be expected to have *P* < 0.05 by chance because of the randomisation.

**Table 1.** Baseline and on-treatment characteristics along with SVR12 rate. For continuous variables, the mean ± standard deviation is presented, while for categorical variables, frequency (percentage) is provided.

|  | All | Received ribavirin |  |  |
| --- | --- | --- | --- | --- |
|  |  | Yes | No | *P* |
|  | 196 (100.0%) | 98 (50.0%) | 98 (50.0%) |  |
| **Baseline factors** |  |  |  |  |
| Randomized under |  |  |  |  |
| First protocol | 133 (67.9%) | 66 (67.3%) | 67 (68.4%) | 0.88 |
| Second protocol | 63 (32.1%) | 32 (32.7%) | 31 (31.6%) |  |
| Days of first line DAA | 45.5 ± 11.5 | 45.5 ± 11.6 | 45.6 ± 11.3 | 0.96 |
| Age (years) | 45.7 ± 10.5 | 45.1 ± 9.7 | 46.3 ± 11.3 | 0.41 |
| Female | 61 (31.1%) | 34 (34.7%) | 27 (27.6%) | 0.28 |
| BMI (kg/m<sup>2</sup>) | 25.4 ± 4.6 | 24.3 ± 3.7 | 26.5 ± 5.0 | <0.001 |
| HCV viral load (log<sub>10</sub> IU/mL) | 5.7 ± 0.9 | 5.6 ± 0.9 | 5.8 ± 0.9 | 0.09 |
| HCV genotype |  |  |  |  |
| 1a | 163 (83.2%) | 83 (84.7%) | 80 (81.6%) | 0.57 |
| 1b | 33 (16.8%) | 15 (15.3%) | 18 (18.4%) |  |
| IL28 genotype |  |  |  |  |
| CC | 59 (31.4%) | 28 (29.8%) | 31 (33.0%) | 0.64 |
| CT or TT | 129 (68.6%) | 66 (70.2%) | 63 (67.0%) |  |
| Fibroscan result (kPa) | 5.0 ± 1.0 | 5.0 ± 1.0 | 5.0 ± 1.1 | 0.93 |
| Albumin (g/L) | 44.1 ± 3.8 | 44.4 ± 3.7 | 43.9 ± 3.8 | 0.38 |
| Bilirubin (umol/L) | 9.9 ± 6.0 | 10.1 ± 6.7 | 9.7 ± 5.3 | 0.68 |
| Creatinine (umol/L) | 75.8 ± 15.6 | 74.5 ± 15.1 | 77.1 ± 16.1 | 0.25 |
| ALT (U/L) | 83.7 ± 94.9 | 81.6 ± 81.1 | 85.8 ± 107.3 | 0.76 |
| AST (U/L) | 50.7 ± 35.8 | 50.7 ± 34.3 | 50.7 ± 37.4 | 1 |
| ALP (U/L) | 75.4 ± 23.8 | 78.7 ± 24.8 | 72.0 ± 22.5 | 0.05 |
| HIV infection | 67 (34.2%) | 34 (34.7%) | 33 (33.7%) | 0.88 |
| Current depression | 38 (19.4%) | 20 (20.4%) | 18 (18.4%) | 0.72 |
| Current illicit substance use | 29 (14.8%) | 13 (13.3%) | 16 (16.3%) | 0.55 |
| **On-treatment factors** |  |  |  |  |
| HCV viral load (log<sub>10</sub> IU/mL) |  |  |  |  |
| Day 3 | 2.4 ± 0.7 | 2.4 ± 0.7 | 2.5 ± 0.7 | 0.78 |
| Day 7 | 1.9 ± 0.6 | 1.9 ± 0.6 | 1.9 ± 0.6 | 0.84 |
| Day 14 | 1.5 ± 0.4 | 1.5 ± 0.4 | 1.6 ± 0.4 | 0.12 |
| Day 0-3 absolute decline | 3.3 ± 0.7 | 3.3 ± 0.8 | 3.4 ± 0.7 | 0.19 |
| Day 0-3 relative decline (%) | 57.7 ± 11 | 57.3 ± 11.0 | 58.2 ± 10.0 | 0.57 |
| SVR12 | 134 (69.4%) | 68 (70.8%) | 66 (68.0%) | 0.674 |

On-treatment virological responses were similar between groups. Mean HCV viral load levels at day 3 (2.4 vs 2.4 log_10_ IU/mL in ribavirin vs no-ribavirin, respectively, *P* = 0.78), day 7 (1.9 vs 1.9 log_10_ IU/mL, respectively, *P* = 0.84), and day 14 (1.5 vs 1.6 log_10_ IU/mL, respectively, *P* = 0.12) did not differ significantly by ribavirin use. Early viral kinetics were likewise comparable: day 0-3 absolute decline (3.3 vs 3.4 log_10_ IU/mL, respectively, *P* = 0.19) and day 0-3 relative decline (57.3 vs 58.2, *P* = 0.57) showed no meaningful differences.

Overall, 62 participants experienced first-line treatment failure. A higher SVR12 was observed in the ribavirin group (70.8% vs 68.0% in the no-ribavirin group), but this difference did not reach statistical significance (*P* = 0.67).

### Viral genome sequencing overview

Samples with detectable viremia underwent deep sequencing at three study time points: baseline (n = 191), day 3 of treatment (n = 25; restricted to those with HCV VL > 500IU/mL), and post-treatment failure (n = 47) (**Figure 1**). Baseline sequencing coverage was therefore available for the majority of participants, with progressively fewer samples sequenced at later time points due to viral suppression during therapy.

**Figure 1.**
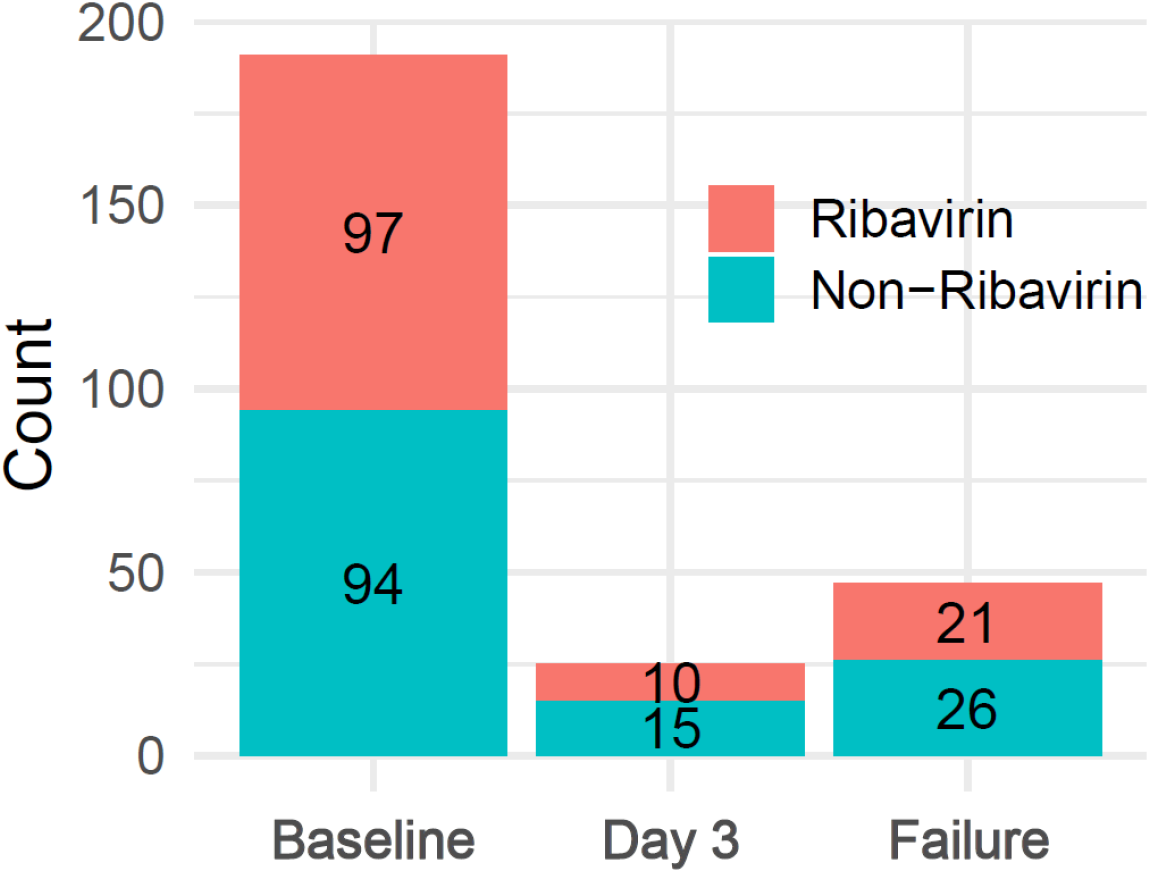
Sequenced participants. Number of plasma samples sequenced at baseline, day 3, and post-treatment failure stratified by ribavirin treatment status.

### Sequencing depth does not confound ribavirin-associated mutation patterns

To account for variation in sequencing depth across samples and time points, we evaluated whether depth could bias ribavirin-associated mutation estimates by comparing models with and without depth adjustment. Including coverage depth as a covariate did not alter the estimated ribavirin effects (average Δ-estimate = 0.04 log-odds), and depth itself was not associated with mutation frequency (*P* = 0.63). Model fit was unchanged with depth included, indicating that sequencing coverage had a negligible influence on ribavirin versus non-ribavirin comparisons at any time point. Therefore, we only considered models not adjusting for depth in subsequent analyses.

### Impact of ribavirin on viral mutagenesis and quasispecies diversity

While there was no evidence of differences in mutational profiles between the ribavirin and no-ribavirin groups at baseline (*P* = 0.12) and day 3 (*P* = 0.98), ribavirin treatment was associated with a significant increase in mutational frequency at post-treatment failure (*P* = 0.0065, **Figure 2A**).

**Figure 2.**
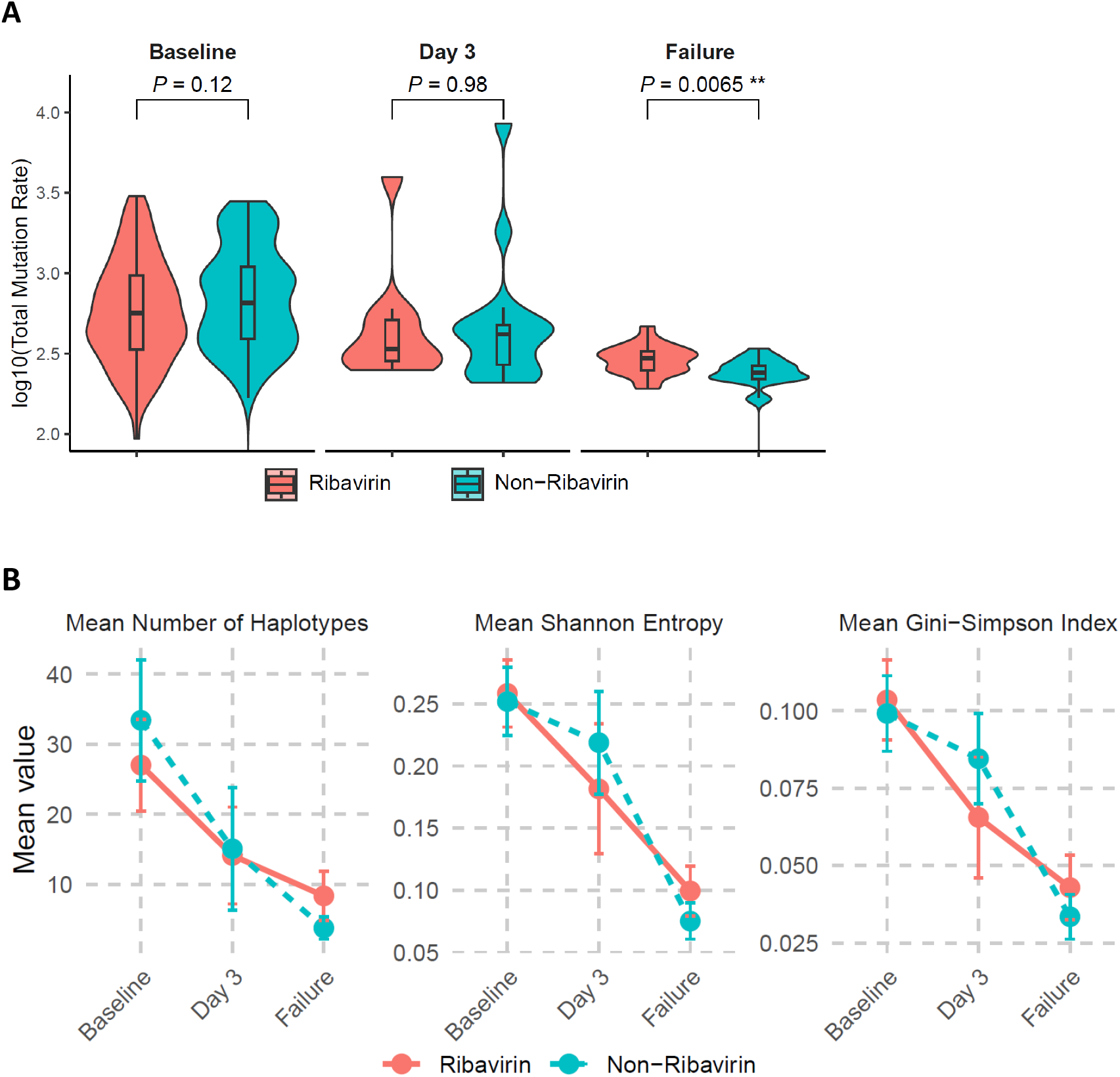
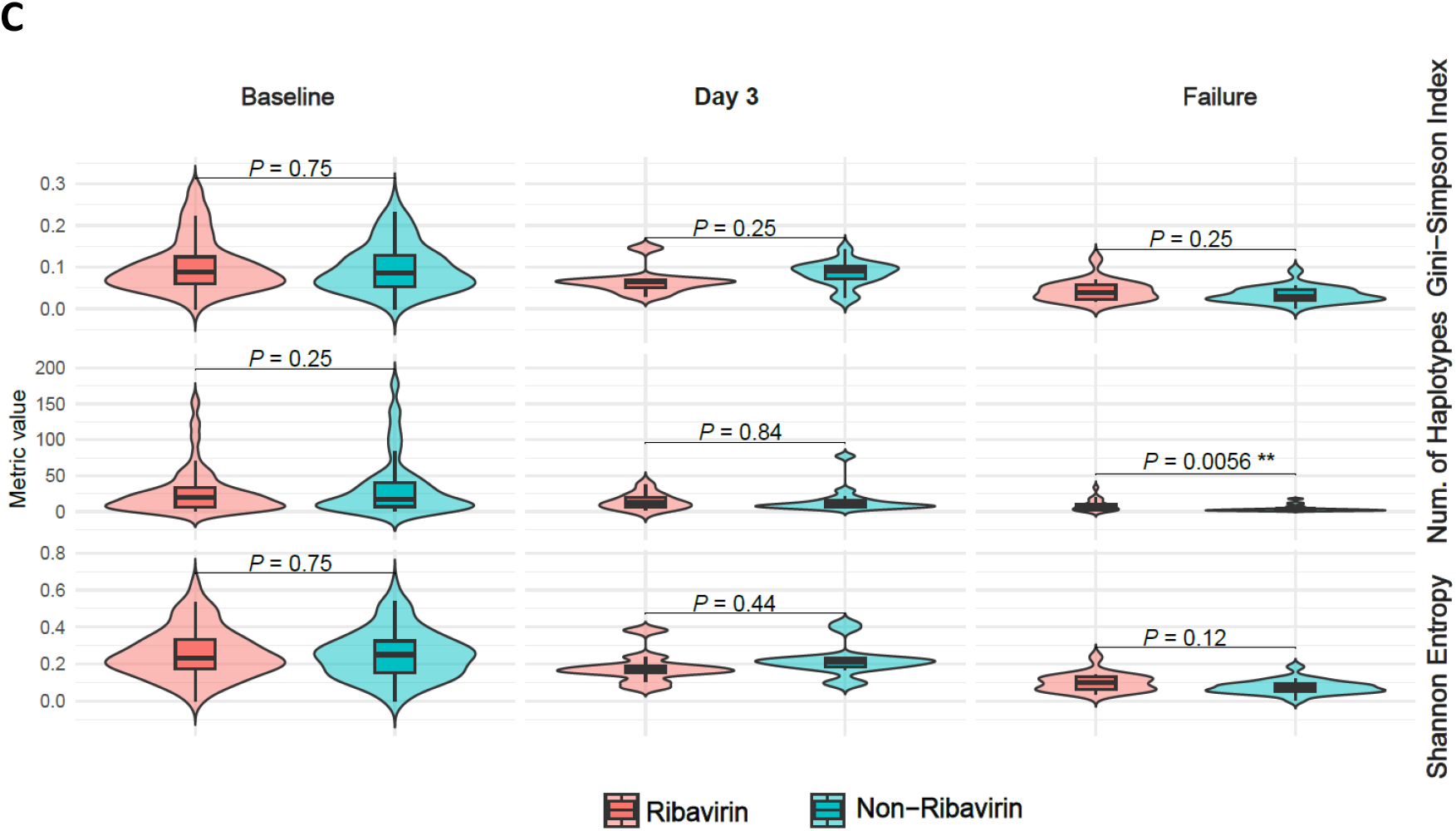
Impact of ribavirin on viral mutagenesis and quasispecies diversity. (A) Normalized mutation rate across baseline, day 3, and post-treatment failure, calculated as the number of mutations per sample divided by the number of genomic positions with ≥100× sequencing depth, thereby accounting for differences in genome coverage between samples. (B) Temporal changes in quasispecies diversity assessed by number of haplotypes, Shannon entropy, and Gini-Simpson index, with 95% confidence intervals shown. (C) Diversity metrics. Data are shown as raw values. Statistical comparisons were performed using linear models adjusting for sequencing depth (log-transformed coverage). P-values are adjusted for multiple testing (Benjamini-Hochberg correction) (* *P* < 0.05; ** *P* < 0.01).

Quasispecies diversity, measured using number of haplotypes, Shannon entropy (sequence variability), and the Gini-Simpson index (probability of sampling distinct viral variants), declined from baseline to day 3 and to failure in both treatment groups, consistent with the suppressive effect of DAA therapy on viral heterogeneity. Diversity estimates were adjusted for sequencing coverage and statistical comparisons were corrected for multiple testing. Overall, no significant differences between treatment groups were observed at baseline or day 3 across any diversity metric. At treatment failure, however, the number of haplotypes was significantly higher in the ribavirin group (adjusted *P* = 0.0056), indicating increased viral diversity in these patients, potentially consistent with a modest increase in mutagenic activity among those who failed therapy. No statistically significant differences were observed for Shannon entropy or the Gini-Simpson index after multiple testing correction (**Figure 2B-C**).

Taken together, these results highlight that while DAA therapy rapidly reduces overall viral diversity, ribavirin selectively enhances mutation accumulation and subtly reshapes quasispecies structure later in treatment.

### Ribavirin induces a broad mutational spectrum

**Figure 3A** illustrates the types of mutations detected in the viral genome. We observed clear enrichment of canonical ribavirin-associated transitions, with G→A substitutions showing increased odds under ribavirin exposure (odds ratio (OR) = 1.14, *P* = 0.026), alongside C→T transitions (OR = 1.15, *P*= 0.004). Additional substitution types also exhibited significantly higher odds in the ribavirin group, including A→G (OR = 1.19, *P* = 0.005), T→C (OR = 1.19, *P* = 0.023), C→G (OR = 1.21, *P* = 0.010), and T→A (OR = 1.28, *P* = 0.026).

**figure 3.**
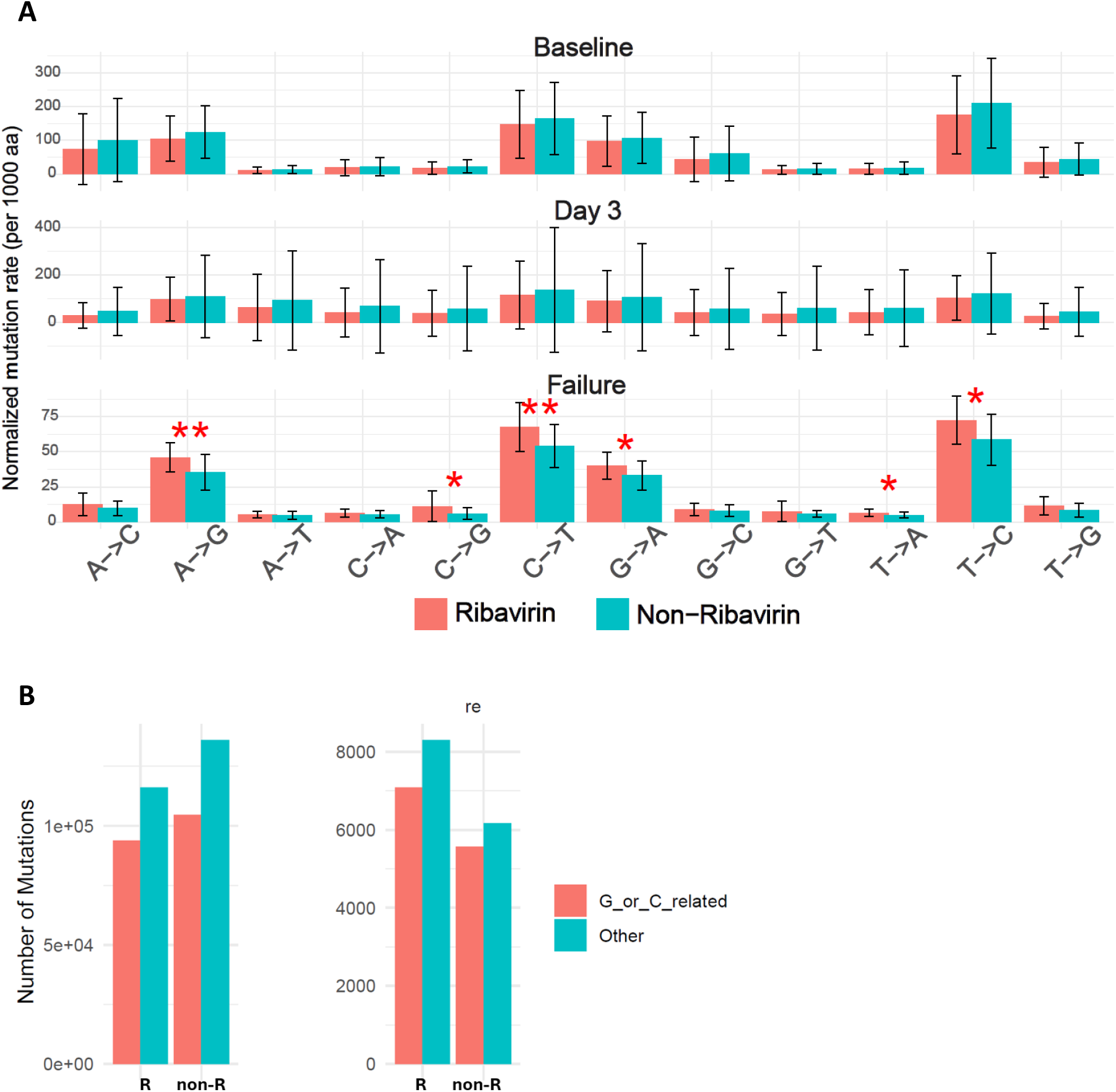
impact of ribavirin on mutational spectrum. (a) distribution of base substitution types observed across the hcv genome (* *p* < 0.05; ** *p* < 0.01). (b) comparison of substitutions away from guanosine between ribavirin and no-ribavirin groups, including analysis of the reverse complement to control for strand orientation. no significant enrichment of g-related substitutions was observed (wilcoxon rank-sum test, all *p* > 0.05).

To test the hypothesis that ribavirin induces guanosine-targeted mutagenesis, we specifically quantified nucleotide substitutions involving guanosine (G) bases (e.g., G→A changes). Because HCV is a positive-strand RNA virus and substitutions can appear on either strand depending on alignment, we repeated the analysis on the reverse complement to ensure strand-independent interpretation. We then tested whether G-related substitutions were enriched in the ribavirin-treated group compared to controls. As shown in **Figure 3B**, we found no evidence of enrichment of G-related substitutions in the ribavirin group.

### Region-specific distribution of ribavirin-associated mutations

To determine whether ribavirin-associated mutagenesis was uniformly distributed across the HCV genome or concentrated within specific regions, we quantified region-specific mutation rates across all major viral genes at baseline, day 3 of treatment, and post-treatment failure (**Figure 4**).

**Figure 4.**
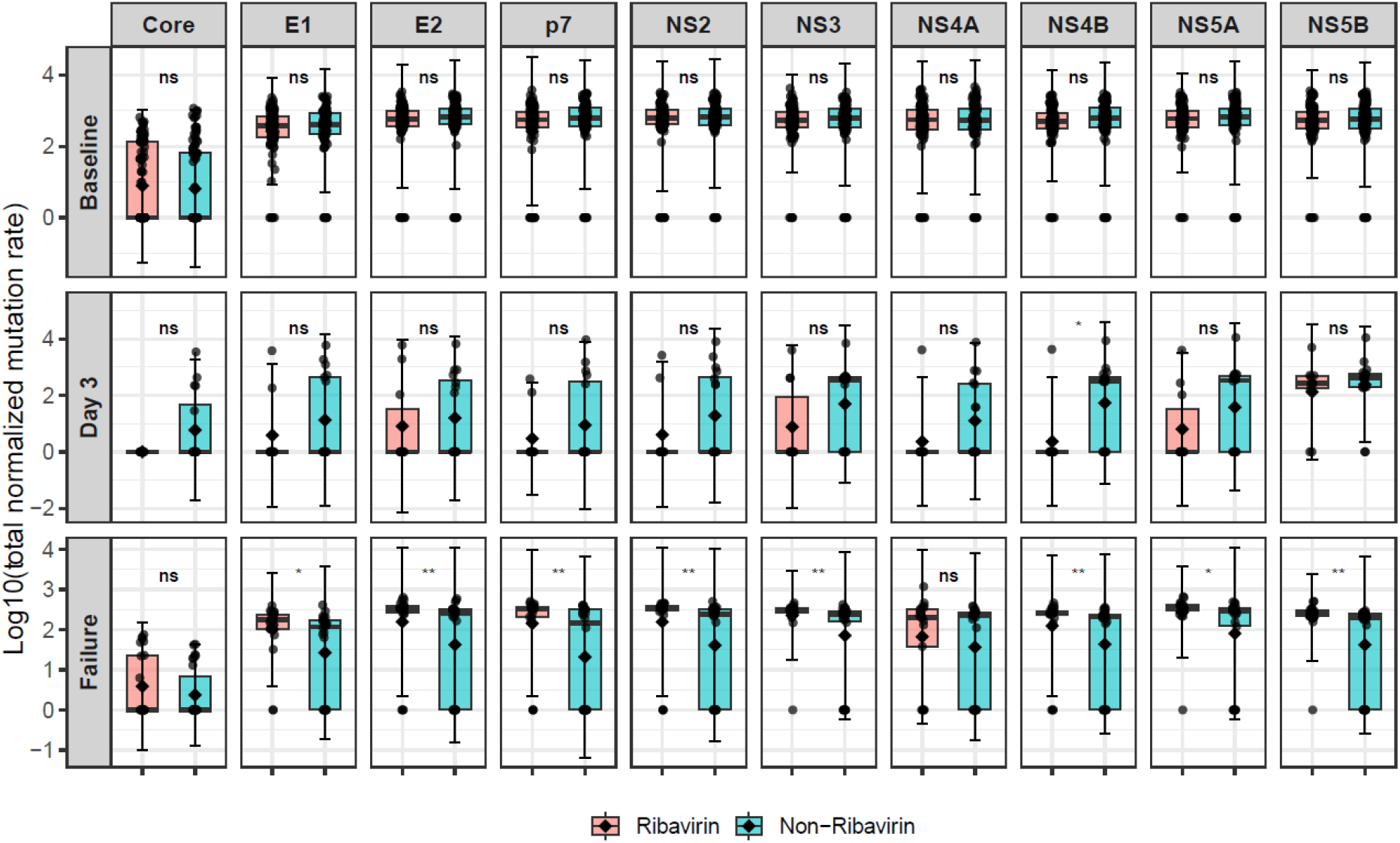
Region-specific mutation rates. Normalized mutation rates across major HCV genomic regions across time points are stratified by ribavirin exposure (* *P* < 0.05; ** *P* < 0.01).

At baseline, there was no evidence that mutation rates differed between ribavirin and no-ribavirin groups in any region (all *P* > 0.17), indicating comparable pre-treatment mutational landscapes, as expected from the randomisation.

At day 3, regional mutation rates were more variable; however, interpretation was limited by small sample numbers and incomplete genome recovery. Only NS4B showed a significantly higher mutation rate in the ribavirin group (*P* = 0.028), with no evidence of differences in other regions.

In contrast, at post-treatment failure, ribavirin exposure was associated with significantly higher mutation rates across most viral regions, including E1, E2, p7, NS2, NS3, NS4B, NS5A, and NS5B (*P* = 0.044, 0.008, 0.003, 0.003, 0.006, 0.008, 0.026, and 0.003, respectively), while there was no evidence of differences in Core (*P* = 0.26) or NS4A (*P* = 0.53). These findings indicate that ribavirin-associated mutagenesis in replicating viruses is broadly distributed across both structural and non-structural genes rather than confined to specific genomic loci.

### Selective pressure dynamics

The temporal changes in the ratio of non-synonymous to synonymous mutations (dN/dS), reflecting the balance between amino acid-altering (non-synonymous, NS) and silent (synonymous, S) substitutions, across the two treatment groups are shown in **Figure 5**. An increase was observed at day 3 (*P* = 5.5e-5), followed by a marked decline at treatment failure (*P* = 8.5e-7). Ribavirin was associated with a trend toward higher dN/dS ratios at day 3 (*P* = 0.06), whereas no significant difference was observed at treatment failure (*P* = 0.11); however, across all time points, mean dN/dS values remained below one, consistent with predominant purifying selection. To assess whether synonymous mutations contributed disproportionately to these patterns, we examined the distribution of mutations across codon positions. No enrichment of mutations at third codon positions was observed in the ribavirin group at any time point (all *P* > 0.1), indicating that the observed dN/dS trends are unlikely to be driven by an increased rate of synonymous substitutions.

**Figure 5.**
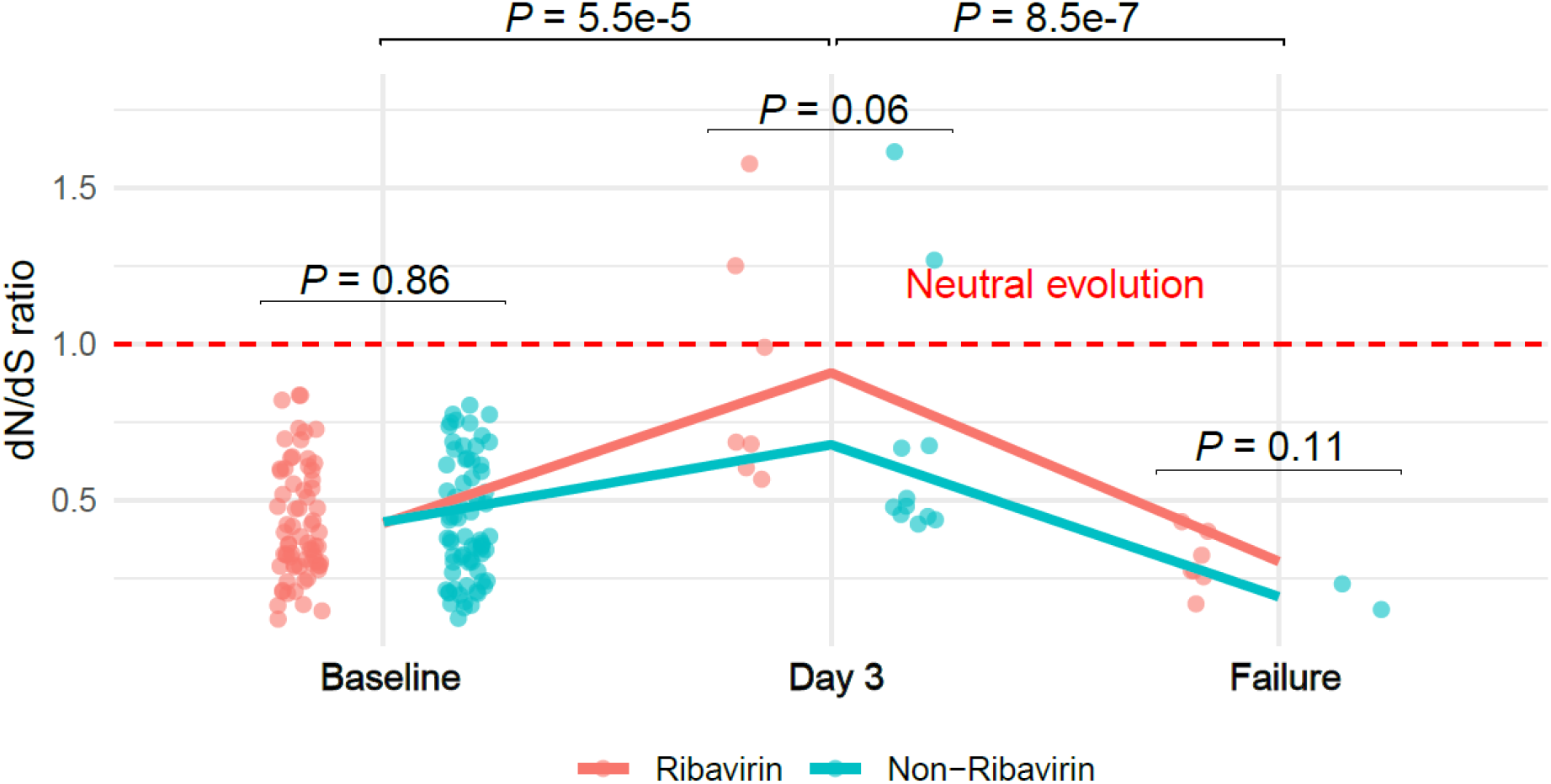
Dynamics of selective pressure during ribavirin treatment. Temporal trends in dN/dS across baseline, day 3, and post-treatment failure. Each point represents a single sample (patient-level dN/dS estimate).

## Discussion

In the era of interferon-based therapy, ribavirin improved treatment outcomes (Poynard et al., 1998). With advances in DAAs, ribavirin is no longer routinely used; however, interest remains in its potential role against emerging or re-emerging viral infections. Despite this, clinical evidence explaining its mechanism of action remains limited.

This study provides the most comprehensive clinical evidence on ribavirin’s mutagenic role in patients infected with HCV. Using deep sequencing of serial samples from a large, randomized trial, we show that ribavirin exposure increases viral mutation rates and diversifies HCV quasispecies. The resulting mutational signature was broad, encompassing both canonical (G→A, C→U) and non-canonical substitutions. These patterns are consistent with ribavirin acting as a general mutagen rather than targeting specific nucleotides. The absence of specific enrichment of G-related changes argues against a simple guanosine-analogue-mediated mechanism.

A limitation is that mutations were best detected at treatment failure, biasing detection toward mutations with minimal replication fitness cost. Early treatment samples (day 3) had lower viral loads, limiting the numbers that could be sequenced and sequencing depth. There was limited evidence of increased mutational rates at day 3 in those exposed to ribavirin, with only a slight increase detected in NS4B, consistent with either insufficient exposure to generate detectable mutations or rapid purging of deleterious variants. A transient rise in dN/dS at day 3, but with average dN/dS values remaining below one, indicates most non-synonymous mutations are deleterious and under purifying selection.

Taken together, our results support a mechanism consistent with error catastrophe, with ribavirin inducing a substantially higher mutational burden *in vivo* than observed in short-duration cell-culture models. This may reflect longer clinical exposure, larger effective viral population sizes, and longitudinal sampling capturing gradual mutation accumulation.

We did not find evidence that mutagenesis was focused on specific genomic regions. Ribavirin-associated mutagenesis at treatment failure was broadly distributed across the HCV genome. Increased mutation rates were observed across both structural and non-structural regions (E1/E2, p7, NS2, NS3, NS4B, NS5A, and NS5B), with no focal accumulation within genes that are antiviral drug targets. Detectable mutations likely represent the non-lethal fraction of a larger pool of ribavirin-induced errors, as highly deleterious lineages are likely lost during treatment. The relative sparing of Core and NS4A is consistent with strong functional constraints, whereby mutations in these regions incur a disproportionate fitness cost and are therefore purged, rather than reflecting protection from mutagenesis.

Ribavirin likely disrupts base-pairing during viral RNA synthesis, generating a distributed mutational load. Over time, this accumulation reduces replication competence. Although ribavirin increases the overall mutational burden the virus remains constrained by functional limits, and most induced changes do not confer advantage. This pattern supports an error-catastrophe model rather than adaptive evolution. These findings also explain ribavirin’s clinical benefit even when direct antiviral activity is minimal: by increasing genetic instability, it reduces the likelihood of resistant or rebound variants surviving post-treatment.

There are a number of key limitations to the data presented here relevant to its generalisability. Firstly, our analyses only included HCV genotypes 1a and 1b. Whilst these account for a majority of HCV found globally, effects could be different in other genotypes. Secondly, the study only used a particular adult dose of oral ribavirin, the finding may differ with other dosing approaches and delivery routes (e.g. inhaled ribavirin). Sequence analysis does not allow us to assess all proposed mechanisms of ribavirin action, such as depletion of intracellular GTP via IMPDH inhibition or immunomodulatory effects. Furthermore, sequencing-based analyses are affected by inherent technical limitations, including variable depth across time points, reduced recoverability of viral genomes during periods of low viremia, and the difficulty of distinguishing true minority variants from technical noise. Although we formally evaluated and ruled out sequencing depth as a confounder of mutational patterns, other potential biases, such as PCR-related drop-out or incomplete haplotype recovery, cannot be fully excluded.

Beyond HCV, the implications extend to other RNA viruses. The demonstrated *in vivo* mutagenic potential supports further exploration of ribavirin and related nucleoside analogues as adjuncts in combination therapies against emerging pathogens, particularly where DAAs are limited. Future studies integrating host pharmacogenomics and viral dynamics modelling could refine ribavirin’s optimal therapeutic window and inform rational drug design.

## Data Availability

All data produced in the present study are available upon reasonable request to the authors.

## Acknowledgements

STOPHCV-1 was funded by the National Institutes of Health Research (NIHR) Efficacy and Mechanism Evaluation programme (grant number 14/02/17). MM, AB, and GC are supported in part by the NIHR Biomedical Research Centre of Imperial College Healthcare NHS Trust. ASW is supported by core support to the MRC Clinical Trials Unit at UCL (MC_UU_00004/03). ASW and GC are NIHR Senior Investigators. This work was partly carried out within the BRC-supported Molecular Diagnostics Unit. The views expressed in this publication are those of the authors and not necessarily those of the NHS, the National Institute for Health Research, or the Department of Health.

